# Systematic review of environmental factors associated with severe fever with thrombocytopenia syndrome occurrences

**DOI:** 10.1101/2025.10.01.25337120

**Authors:** Gayoung Lee, Hyun-Kyung Kim, Arata Hidano, Sung-mok Jung

**Affiliations:** Department of Epidemiology, Gillings School of Global Public Health, University of North Carolina at Chapel Hill, North Carolina, USA; Department of Public Health Sciences, Graduate School of Public Health, Seoul National University, Seoul, Republic of Korea; Department of Global Health and Development, Faculty of Public Health and Policy, London School of Hygiene and Tropical Medicine, London, UK; Department of Infectious Disease Epidemiology and Dynamics, Institute of Tropical Medicine, Nagasaki University, Nagasaki, Japan; Saw Swee Hock School of Public Health, National University of Singapore, Singapore; Carolina Population Center, University of North Carolina at Chapel Hill, North Carolina, USA

## Abstract

Severe fever with thrombocytopenia syndrome (SFTS) is an emerging tick-borne viral disease with high fatality among older adults, posing a growing public health threat across Asia with increasing potential for geographic spread beyond the region. Yet control efforts remain largely confined to individual-level tick bite prevention, reflecting persistent gaps in understanding the ecological mechanisms that sustain SFTS transmission, particularly how environmental factors shape transmission dynamics across vectors, animal reservoirs, and human populations. We systematically reviewed 2,910 studies to synthesize evidence on associations between environmental factors and SFTS occurrences across populations. Temperature, humidity, precipitation, elevation, and land cover are consistently linked to human SFTS occurrence through non-linear, often reverse U-shaped, relationships, underscoring the need for analytical frameworks capable of capturing the inherently non-linear nature of environmental influences. However, we identified no quantitative assessments of how environmental factors shape SFTS occurrence in vectors or animal reservoirs, nor any stage-specific assessment of how these factors act across the transmission cycle, leaving multifaceted environmental effects acting across the tick–animal–human interface collapsed into oversimplified estimates based solely on human cases. To address these critical gaps, future research must prioritize stage-specific elucidation of environmental drivers across the SFTS transmission cycle through mechanistic modeling approaches integrated with transboundary surveillance under a One Health framework. As climate and land-use changes continue to reshape vector habitats and expand regions at risk, such efforts will be essential for enhancing ecological understanding and guiding One Health-grounded surveillance and control strategies that can mitigate the growing transboundary burden of SFTS.

## Introduction

Severe fever with thrombocytopenia syndrome (SFTS) is an emerging tick-borne zoonotic disease caused by SFTS virus (SFTSV), posing a growing public health threat across Asia. First identified in China in 2010 [1], the virus has since spread to neighboring countries, including Japan and the Republic of Korea (hereafter “South Korea”), where sustained transmission to humans has been established. Clinically, SFTS manifests as high fever and thrombocytopenia and can progress to severe multi-organ failure. Age is a major factor of disease severity, with case fatality risks rising sharply from 4–9% in younger adults to 25–35% among those aged over 60 years [2]. Reflecting its substantial burden and the absence of accessible vaccines or antiviral treatments [3], the World Health Organization designated SFTS as one of ten priority infectious diseases requiring urgent research [4].

Despite such global recognition of its public health threat, effective control of SFTS remains challenging due to its complex ecological dynamics that sustain viral circulation and facilitate spillover to humans. *Haemaphysalis longicornis* (*H. longicornis*) has been identified as the primary tick vector (while the virus has been occasionally detected in other tick species) [5]; however, its interactions with a wide range of reservoir hosts—including wild and domestic mammals and avian species [6]—remain poorly characterized. This uncertainty has impeded the development of ecologically grounded interventions, leaving current control strategies largely reliant on individual-level preventive measures (e.g., tick bite avoidance such as wearing long sleeves during outdoor activities [7]), which are unlikely to effectively contain the continued geographic expansion of the virus [8]. While human SFTS cases are yet confined to Asia, the detection of *H. longicornis* in the Western Pacific Islands [6] and multiple U.S. states [9] has raised legitimate concerns about the potential emergence of new endemic areas.

In this context, synthesizing evidence on how environmental factors shape SFTS occurrence across vectors, animal hosts, and humans provides critical insights into its transmission dynamics within a broader ecological framework. Climatic and topographic conditions—including temperature, humidity, precipitation, and elevation—are well established as key determinants of tick-borne disease dynamics by regulating the abundance, distribution, and behavior of both vectors and hosts (**Figure 1**) [10]. With accelerating global climate change, these drivers are expected to exert even greater influence, potentially reshaping vector habitats and facilitating the spread of pathogens into previously unaffected regions. However, the utility of such synthesis depends critically on the methodological rigor of individual studies. Existing studies often rely on coarse surveillance data and simplified analytical approaches that fail to capture the inherently dynamic nature (non-linear) of ecological systems. These limitations may bias inferences about how environmental drivers influence transmission risk, particularly for emerging pathogens such as SFTSV, where ecological knowledge remains limited. To address these challenges, we conducted a systematic review of the literature examining associations between environmental factors and SFTS occurrence across vectors, animal hosts, and human populations, with a focus on the analytic approaches used. This synthesis aims to consolidate current evidence and discuss key research priorities to inform more ecologically grounded surveillance and control strategies for SFTS.

**Figure 1.**
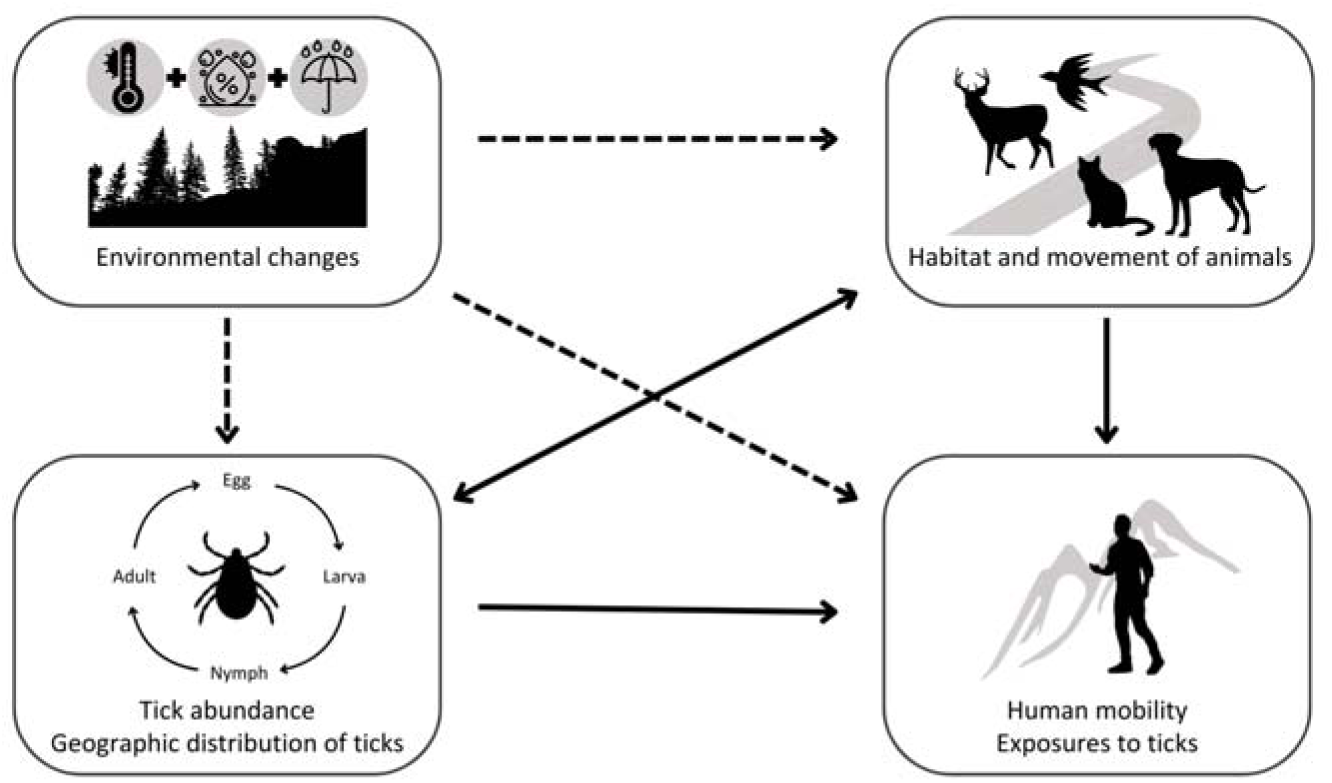
Schematic diagram of the influence of environmental factors on severe fever with thrombocytopenia syndrome transmission. Combined schematic representing the transmission dynamics of SFTSV (solid arrows) and the potential influence of environmental factors (dashed arrows) on tick vectors, animal, and human populations.

## Methods

### Search strategy

We conducted a systematic review following the Preferred Reporting Items for Systematic Reviews and Meta-Analyses guidelines, with a prespecified protocol registered in the International Prospective Register of Systematic Reviews [11]. Our search included articles published in English up to October 31, 2024, across five bibliographic databases: *Embase, Global Health*, *PubMed*, *Scopus*, and *Web of Science*. The search strategy combined terms for the key components: SFTS outcomes and environmental factors (including climatic and topographical variables). No restrictions were applied to specific epidemiological outcomes, allowing for comprehensive inclusion of studies examining environmental influences on SFTS occurrence across vectors (ticks), non-human reservoirs (wildlife and domestic animals), and human populations. Search strings were adapted for the indexing syntax of each database, with full details provided in **Table S1**.

### Study selection

After removing duplicates, studies were screened for eligibility following the pre-defined criteria: (i) original studies using empirical SFTS outcome data (excluding those solely focused on tick distribution), and (ii) studies employing analytic approaches to quantify the relationship between environmental factors and SFTS outcomes beyond simple descriptions (excluding those only examining correlations). Further details on the eligibility criteria are provided in **Table S2**. Screening was conducted in two stages: an initial screening of titles and abstracts, followed by full-text review. Two authors independently performed the screening using the *Rayyan* software. Any discrepancies were resolved through discussion and consensus, with adjudication by senior authors when necessary. To ensure comprehensive coverage, reference lists of included studies and relevant review articles were also manually screened.

### Quality assessment

Two authors independently assessed the risk of bias in all included studies using the JBI Critical Appraisal Tool [12]. This tool evaluates the risk of bias across 10 domains, with each domain rated as low (0), moderate (0.5), or high (1). Minor adaptations were made to address methodological nuances specific to this review, including the resolution of analyzed data and the suitability of applied analytic frameworks. Detailed descriptions of the modified assessment criteria and individual study assessments are provided in **Table S3**.

### Narrative synthesis

Data were extracted from the selected studies and systematically summarized in tables (**Tables 1–3** & **Tables S5–S9**). Extracted information includes study period, geographical location (along with climate zones assigned based on the Trewartha classification [13]), environmental factors, SFTS outcomes, analytic methods, key findings, and time lag (defined as the time interval from environmental exposure to SFTS outcome) specifications. The key findings describe the nature of the relationships between environmental factors and SFTS outcomes. Specifically, where available, we extracted the range or category of environmental variables associated with the highest risk of SFTS outcomes—referred to as the “high-risk range” (or “high-risk point” when only a peak value was reported)—to facilitate interpretation of the overall shape of relationships (skewness) along with the observed exposure ranges. When not explicitly provided in the primary manuscripts, these values were approximated through visual inspection. Studies reporting only univariate results without any covariate adjustment were excluded from the narrative synthesis. All extracted data and interpretations were independently reviewed by all authors, with consensus reached to ensure the accuracy of the synthesis.

**Table 1.**
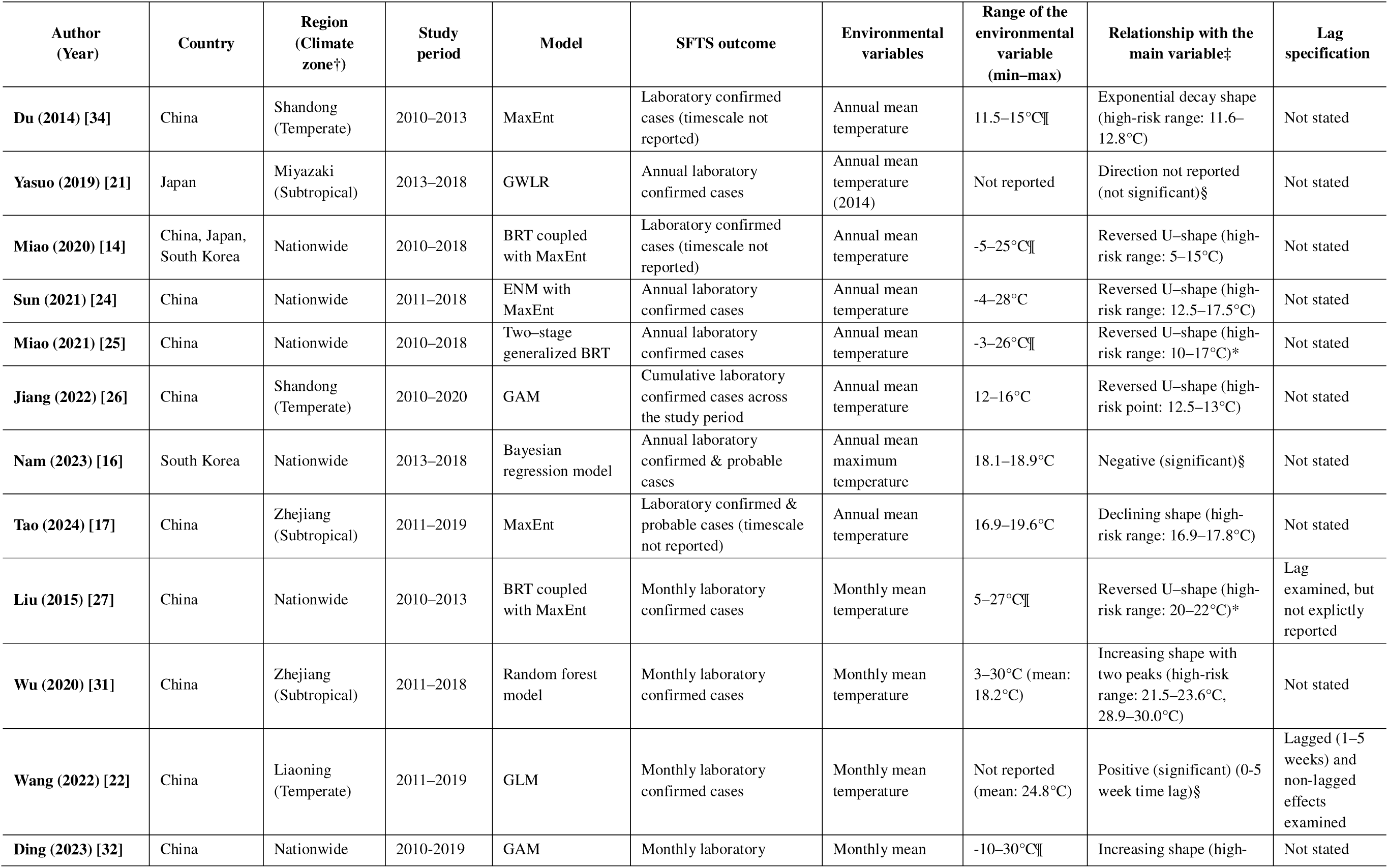
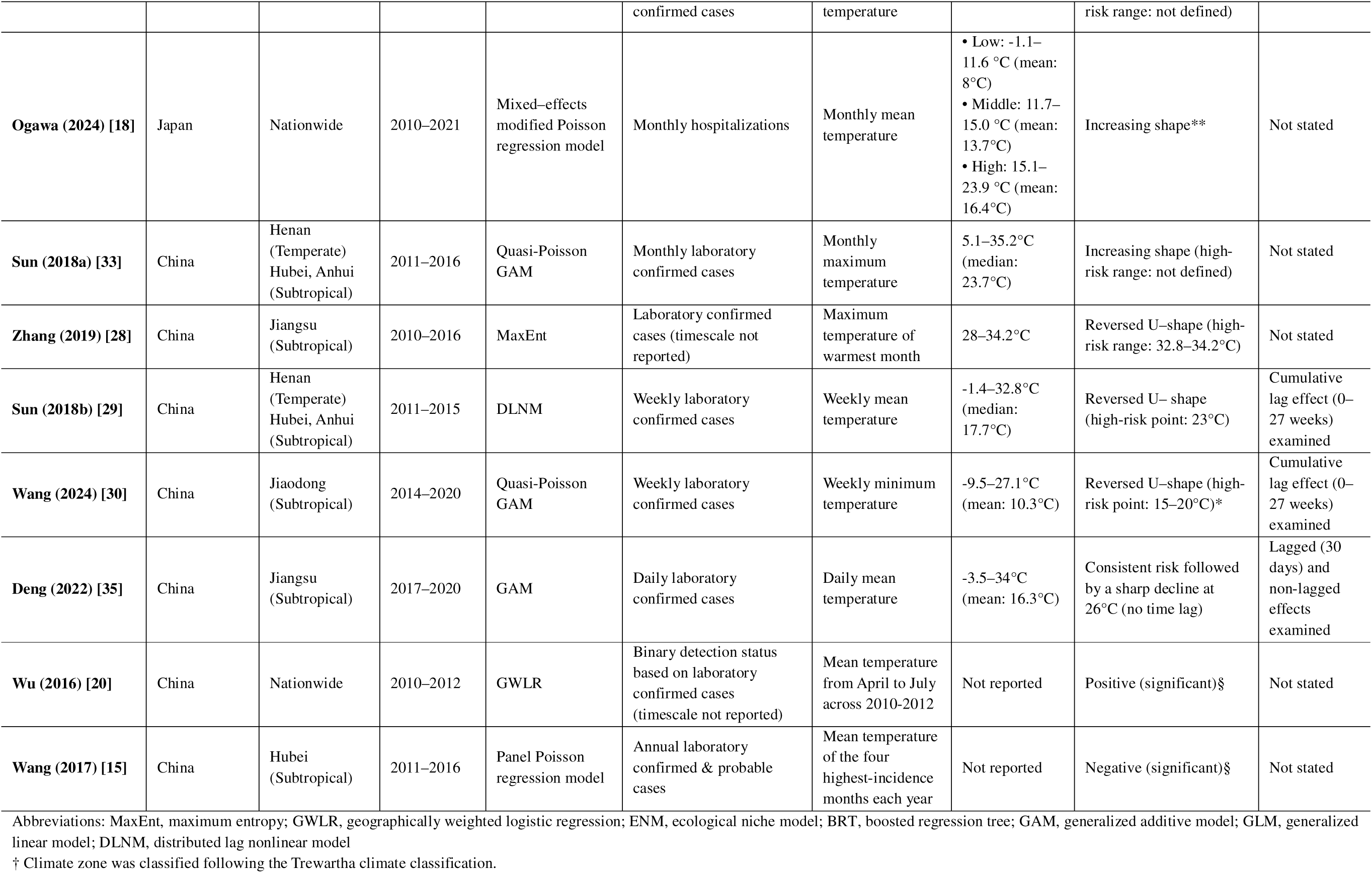
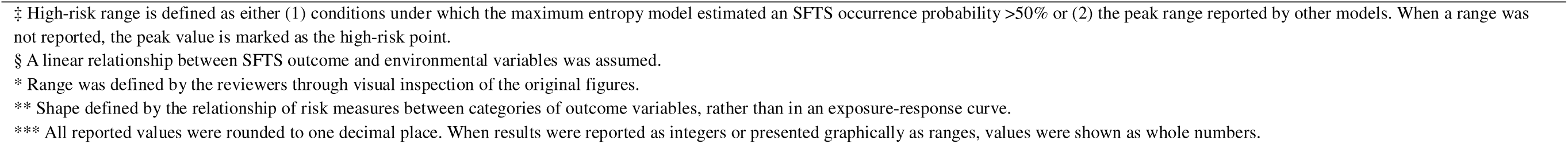
Summary of included papers examining the association between temperature and SFTS occurrence.

**Table 2.** Summary of included papers examining the association between humidity and SFTS occurrence.

| Author (Year) | Country | Region (Climate zone)<sup>†</sup> | Study period | Model | SFTS outcome | Observational variables | Range of the main variable (min–max) | Relationship with the main variable<sup>‡</sup> | Lag specification |
| --- | --- | --- | --- | --- | --- | --- | --- | --- | --- |
| Sun (2021) [24] | China | Nationwide | 2011–2018 | ENM with MaxEnt | Annual laboratory confirmed cases | Annual mean relative humidity | 24.3–91.4% | Reversed U-shape (high-risk range: 63–82%) | Not stated |
| Nam (2023) [16] | South Korea | Nationwide | 2013–2018 | Bayesian regression model | Annual laboratory confirmed & probable cases | Annual mean relative humidity | 66.1–68.8% | Negative (not significant)<sup>§</sup> | Not stated |
| Liu (2015) [27] | China | Nationwide | 2010–2013 | BRT coupled with MaxEnt | Monthly laboratory confirmed cases | Monthly mean relative humidity | 30–85%<sup>¶</sup> | Reversed U-shape (high-risk range 72–75%)* | Lag examined, but not explicitly reported |
| Sun (2018a) [33] | China | Henan (Temperate)<br>Hubei, Anhui (Subtropical) | 2011–2016 | Quasi-Poisson GAM | Monthly laboratory confirmed cases | Monthly mean relative humidity | 51.9–86.6% (median: 72.5%) | Increasing shape (high-risk range: not defined) | Not stated |
| Wu (2020) [31] | China | Zhejiang (Subtropical) | 2011–2018 | Random forest model | Monthly laboratory confirmed cases | Monthly mean relative humidity | 60–89% (mean: 75.4%) | Increasing shape (high-risk range: 81.4–89%) | Not stated |
| Wang (2022) [22] | China | Liaoning (Temperate) | 2011–2019 | GLM | Monthly laboratory confirmed cases | Monthly mean relative humidity | Not reported (mean: 77.4%) | Positive (significant) (0-5 week time lag)<sup>§</sup> | Lagged (1–5 weeks) and non-lagged effects examined |
| Ding (2023) [32] | China | Nationwide | 2010–2019 | GAM | Monthly laboratory confirmed cases | Monthly mean relative humidity | 30–100%<sup>¶</sup> | U-shape followed by plateau (high-risk range: 50–100%)* | Not stated |
| Wang (2024) [30] | China | Jiaodong (Subtropical) | 2014–2020 | Quasi-Poisson GAM | Weekly laboratory confirmed cases | Weekly mean relative humidity | 34.8–87.6% (mean: 66.6%) | Increasing shape (high-risk range: not defined) | Cumulative lag effect (0–27 weeks) examined |
| Deng (2022) [35] | China | Jiangsu (Subtropical) | 2017–2020 | GAM | Estimated tick-to-human transmissibility | Daily mean relative humidity | 31–98% | Decreasing shape (30-day time lag) | Lagged (30 days) and non-lagged effects examined |
| Wu (2016) [20] | China | Nationwide | 2010–2012 | GWLR | Binary detection status based on laboratory confirmed cases (timescale not reported) | Mean relative humidity from April to July across 2010–2012 | Not reported | Positive (significant)<sup>§</sup> | Not stated |
| Wang (2017) [15] | China | Hubei | 2011–2016 | Panel Poisson | Annual laboratory | Mean relative | Not reported | Positive (significant)<sup>§</sup> | Not stated |
|  |  | (Subtropical) |  | regression model | confirmed & probable cases | humidity of the four highest-incidence months each year |  |  |  |

Abbreviations: ENM, ecological niche model; MaxEnt, maximum entropy; BRT, boosted regression tree; GAM, generalized additive model; GLM, generalized linear model; GWLR, geographically weighted logistic regression.
† Climate zone was classified following the Trewartha climate classification.
‡ High-risk range is defined as either (1) conditions under which the maximum entropy model estimated an SFTS occurrence probability >50% or (2) the peak range reported by other models. When a range was not reported, the peak value is marked as the high-risk point.
§ A linear relationship between SFTS outcome and environmental variables was assumed.
¶ Range of the main variable was defined by the authors using the graph's X-axis.
\* Range was defined by the reviewers through visual inspection of the original figures.
\*\* All reported values were rounded to one decimal place. When results were reported as integers or presented graphically as ranges, values were shown as whole numbers.

**Table 3.** Summary of included papers examining the association between precipitation and SFTS occurrence.

| Author (Year) | Country | Region (Climate zone†) | Study period | Model | SFTS outcome | Observational variables | Range of the main variable (min–max) | Relationship with the main variable‡ | Lag specification |
| --- | --- | --- | --- | --- | --- | --- | --- | --- | --- |
| Miao (2021) [25] | China | Nationwide | 2010–2018 | Two-stage generalized BRT | Annual laboratory confirmed cases | Annual cumulative precipitation in driest quarter | 0–420mm¶ | Increasing shape with a plateau at 80 mm* | Not stated |
| Sun (2021) [24] | China | Nationwide | 2011–2018 | ENM with MaxEnt | Annual laboratory confirmed cases | Annual cumulative precipitation | 286.5–2244.9mm | Reversed U-shape (high-risk range: 700–2250 mm) | Not stated |
| Duan (2023) [36] | China | Shandong (Temperate) | 2010–2021 | MaxEnt | Laboratory confirmed cases (timescale not reported) | Annual cumulative precipitation | 400–1500mm¶ | Reversed U-shape (high-risk range: 743.9–956.1 mm) | Not stated |
| Nam (2023) [16] | South Korea | Nationwide | 2013–2018 | Bayesian regression model | Annual laboratory confirmed & probable cases | Annual cumulative precipitation | 934–1402mm | Positive (not significant)§ | Not stated |
| Liu (2015) [27] | China | Nationwide | 2010–2013 | BRT coupled with MaxEnt | Monthly laboratory confirmed cases | Monthly cumulative precipitation | 0–1750mm | Stepwise increasing shape (inflection at 600 mm)* | Lag examined, but not explicitly reported |
| Ogawa (2024) [18] | Japan | Nationwide | 2010–2021 | Mixed-effects modified Poisson regression model | Monthly hospitalizations | Monthly cumulative precipitation | • Low: 675–1405mm (mean: 1217mm)<br>• Middle: 1406–1851mm (mean: 1608mm)<br>• High: 1852–3946mm (mean: 2331mm) | U-shape (not significant)** | Not stated |
| Wu (2020) [31] | China | Zhejiang (Subtropical) | 2011–2018 | Random forest model | Monthly laboratory confirmed cases | Monthly cumulative precipitation | 16–326mm (mean: 138.9mm) | Bimodal shape (high-risk range: 71.9–90.4mm, 189.4–232.7mm) | Not stated |
| Jiang (2022) [26] | China | Shandong (Temperate) | 2010–2020 | GAM | Total number of laboratory confirmed cases across the study period | Annual mean precipitation | 400–750mm | Reversed U-shape (high-risk point: 650 mm) | Not stated |
| Du (2014) [34] | China | Shandong | 2010–2013 | MaxEnt | Laboratory confirmed | January mean | 2–12 mm | January mean | Not stated |
|  |  | (Temperate) |  |  | cases (timescale not reported) | precipitation, Annual mean precipitation | (January mean precipitation)¶ | precipitation: Reversed U-shape (high-risk range: 5.5–7.8mm), Annual mean precipitation: Not reported (high-risk range: 640–660mm) |  |
| **Wu (2016) [20]** | China | Nationwide | 2010–2012 | GWLR | Binary detection status based on laboratory confirmed cases (timescale not reported) | Mean precipitation from April to July across 2010–2012 | Not reported | Negative (significant)§ | Not stated |
| **Wang (2022) [22]** | China | Liaoning (Temperate) | 2011–2019 | GLM | Monthly laboratory confirmed cases | Monthly mean precipitation | Not reported | Positive (significant) (0–5 week time lag)§ | Lagged (1–5 weeks) and non-lagged effects examined |
| **Ding (2023) [32]** | China | Nationwide | 2010–2019 | GAM | Monthly laboratory confirmed cases | Monthly mean precipitation | 0–1000mm¶ | Constant (high-risk range: not defined) | Not stated |
| **Miao (2020) [14]** | China, South Korea, and Japan | Nationwide | 2010–2018 | BRT coupled with MaxEnt | Laboratory confirmed cases (timescale not reported) | Cumulative precipitation of warmest quarter, cumulative precipitation of coldest quarter | 10–100mm (warmest quarter)¶<br>0–60mm (coldest quarter)¶ | • Logistic shape (warmest quarter) (high-risk range: 60–100mm)*<br>• Reversed U-shape (coldest quarter) (high-risk range: 5–20mm)* | Not stated |
| **Tao (2024) [17]** | China | Zhejiang (Subtropical) | 2011–2019 | MaxEnt | Laboratory confirmed & probable cases (timescale not reported) | Cumulative precipitation of wettest month | 131.9–351.9mm | Declining shape (high-risk range: 131.9–224.4mm) | Not stated |

Abbreviations: BRT, boosted regression tree; ENM, ecological niche model; MaxEnt, maximum entropy; GAM, generalized additive model; GWLR, geographically weighted logistic regression; GLM, generalized linear model.
† Climate zone was classified following the Trewartha climate classification.
‡ High-risk range is defined as either (1) conditions under which the maximum entropy model estimated an SFTS occurrence probability >50% or (2) the peak range reported by other models. When a range was not reported, the peak value is marked as the high-risk point.
§ A linear relationship between SFTS outcome and environmental variables was assumed.
¶ Range of the main variable was defined by the authors using the graph's X-axis.
\* Range was defined by the reviewers through visual inspection of the original figures.
\*\* Shape defined by the relationship of risk measures between categories of outcome variables, rather than in an exposure-response curve.
\*\*\* All reported values were rounded to one decimal place. When results were reported as integers or presented graphically as ranges, values were shown as whole numbers

## Results

### Summary of the included papers for review

Following the screening of 2,910 papers, a total of 23 studies were included in the final synthesis to examine the associations between environmental factors and SFTS occurrence (**Figures 2 & 3**). The majority (22 out of 23) analyzed data from a single country, predominantly China (n=19), followed by Japan (n=2) and South Korea (n=1). One multi-country study covered China, Japan, and South Korea [14]. Of the single-country studies, 15 were conducted at the subnational levels, mainly in historically SFTS endemic regions (e.g., Shandong, Hubei, and Zhejiang).

**Figure 2.**
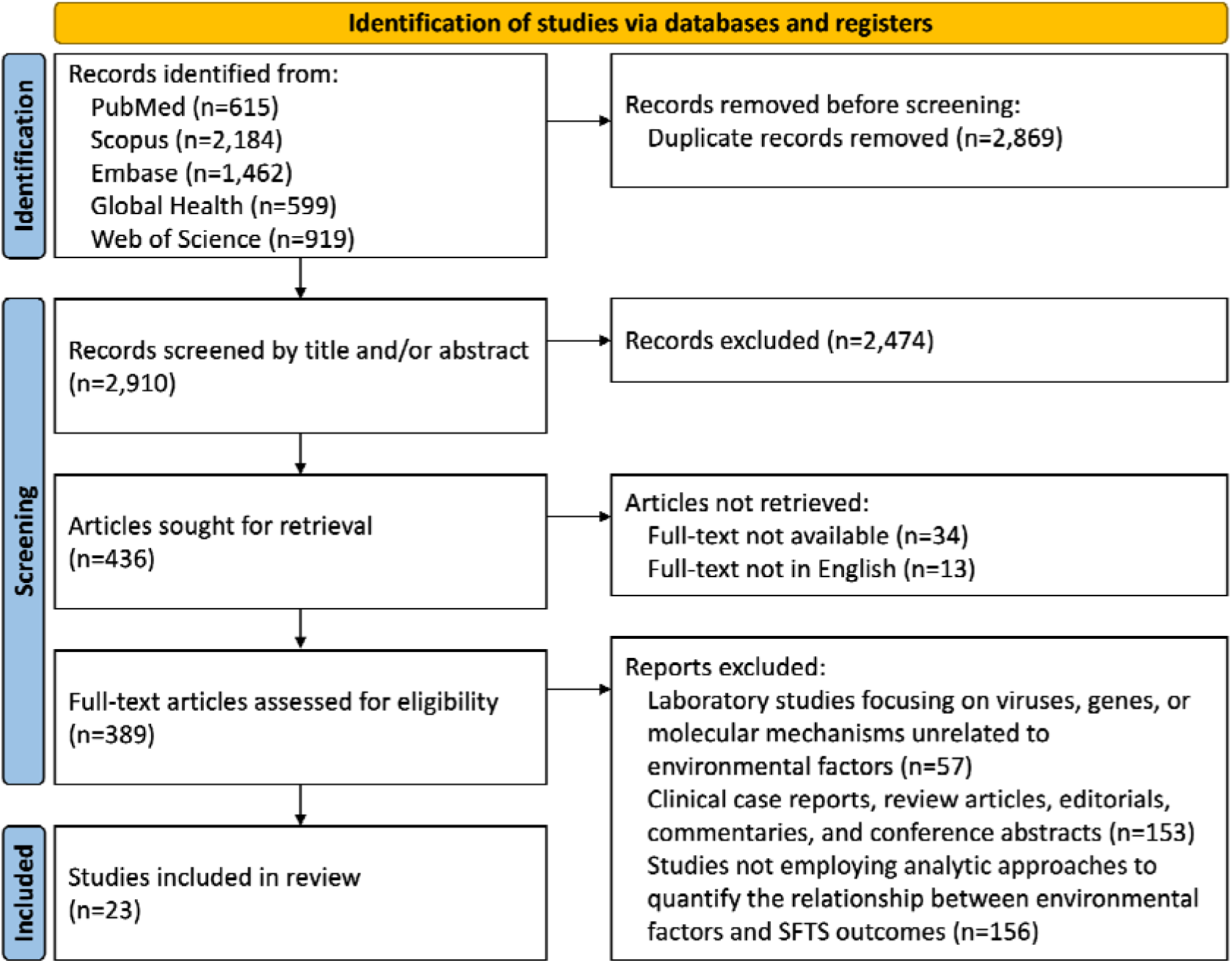
PRISMA flow chart of study selection for a systematic review on severe fever with thrombocytopenia syndrome and environmental factors.

**Figure 3.**
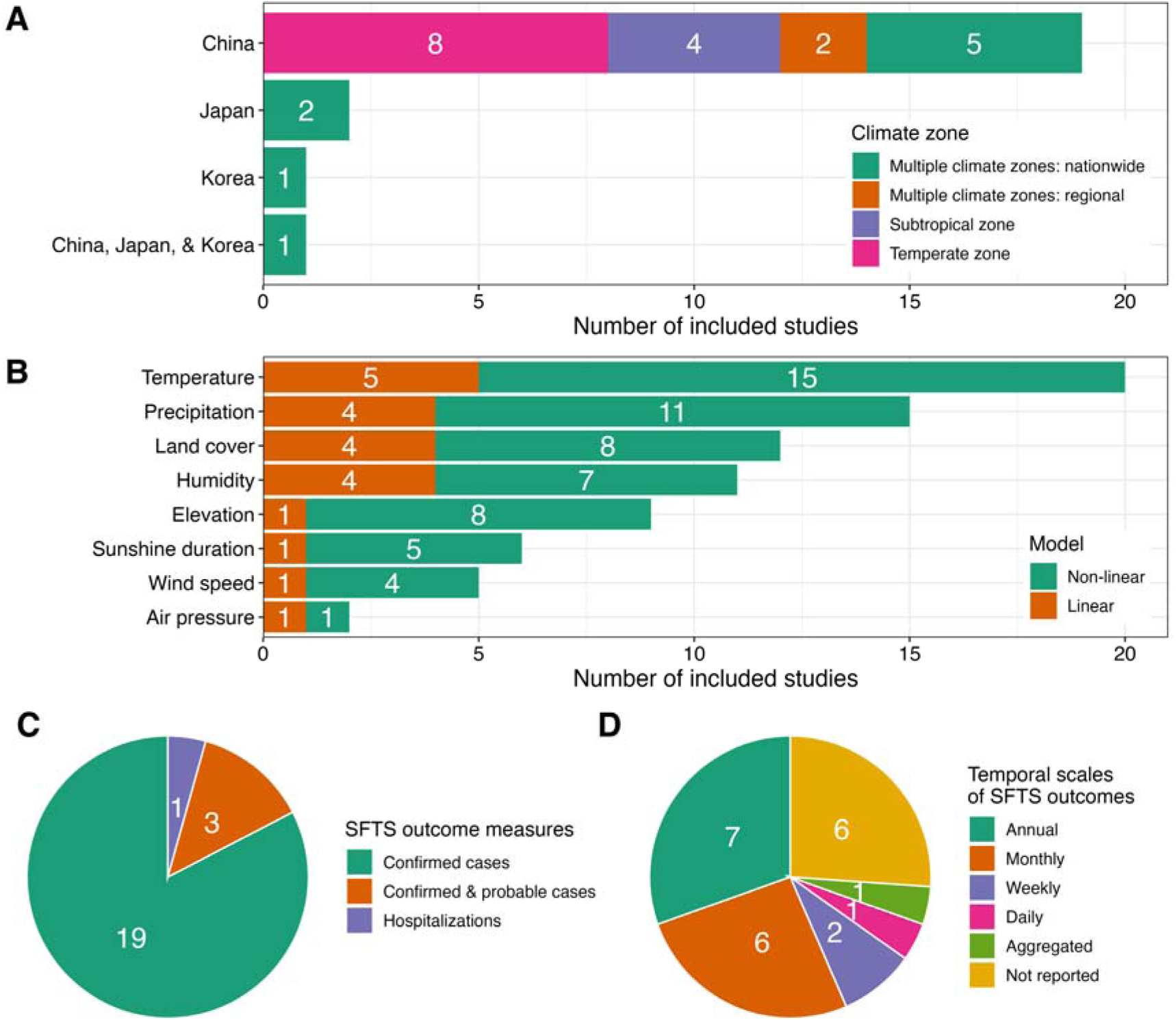
Summary of studies included in a systematic review on severe fever with thrombocytopenia syndrome and environmental factors. (**A**) Number of studies by country and by the climate zone where data were collected. *Multiple climate zones: nationwide* denotes studies conducted at the national level in countries spanning several climate zones under the Trewartha classification, whereas *Multiple climate zones: regional* refers to province-level studies that covered multiple areas across more than one climate zone. (**B)** Number of studies by environmental factors assessed, with classification based on whether linearity was assumed in the analytic approach. (**C**) Proportion of studies by SFTS outcome measures, categorized as laboratory-confirmed cases only, confirmed and probable cases, or hospitalizations. (**D**) Proportion of studies by temporal scales of SFTS outcomes. *Aggregated* refers to studies that applied binary indicators of case occurrence over a defined period longer than one year.

Although our search strategy was designed to capture studies investigating SFTS occurrence across species (i.e., tick vectors, non-human reservoirs, and humans), all studies included focused exclusively on human outcomes. In measuring human SFTS occurrence, all but four studies used laboratory-confirmed cases. Of the exceptions, three included both laboratory-confirmed and probable cases (i.e., clinically diagnosed cases without laboratory confirmation) [15–17], and one analyzed SFTS hospitalizations [18]. SFTS outcomes were reported at varying temporal resolutions, ranging from annual to daily; however, six studies did not specify this information, limiting interpretability and comparability across studies.

Temperature was the most frequently examined environmental factor (n=20), followed by precipitation (n=15), land cover (n=12), humidity (n=11), and elevation (n=9). Other environmental factors explored included sunshine duration (n=6), wind speed (n=5), and air pressure (n=2). Environmental data were most aggregated at either the annual (n=9) or monthly (n=9) level, with each count indicating that at least one variable in the respective studies was measured on that scale. Other studies utilized weekly, or daily scales, or specific summary metrics (e.g., maximum temperature of the warmest month in each year). A detailed summary of environmental variables and their corresponding time scales in each study is provided in **Table S4**.

Most studies (n=16) employed statistical models that allow for potential non-linearity in the relationship between environmental factors and SFTS outcomes, whereas a smaller subset (n=7) applied models constraining the relationship to be linear [15,16,19–23]. Among the non-linear approaches, maximum entropy models were the most frequently used (n=7), followed by generalized additive models (n=5). Time lags were addressed in five studies (**Table S4**). Of these, two studies incorporated lag effects within the analytical framework by applying distributed lag models to estimate cumulative effects over the specified lag periods [28,29]. The remaining three studies examined lag structures through variable definitions, for instance, by including lagged exposures (e.g., temperature with a 1-month lag) as covariates [21,34]; however, one of these did not explicitly report how lagged effects were handled [26].

### Temperature

A total of 20 studies examined the relationship between temperature and human SFTS outcomes (**Table 1**). Of these, 15 accounted for potential non-linearity in the relationship: eight reported a reverse U-shaped relationship (defined as a pattern in which risk increases up to an optimal level followed by a subsequent decline) [14,24–30], four observed an increasing trend [18,31–33], two identified a decreasing trend [17,34], and one reported a pattern of consistent risk followed by a sharp decline at higher temperature [35]. Among studies reporting a reverse U-shape, the high-risk range varied depending on both temporal (annual vs. monthly) and spatial (nationwide vs. subnational) measurement scales.

Nationwide studies using annual mean temperatures typically identified lower high-risk ranges (around 12–15°C [24,25]), whereas those using finer temporal resolutions (monthly mean temperatures) reported higher high-risk ranges (20–22°C [27]). Additionally, a study conducted in a subtropical zone (Zhejiang) reported higher high-risk ranges (17–18°C [17]), whereas two studies in a temperate zone (Shandong) identified lower ranges (around 12–13°C [26,34]). Nationwide studies covering both climate zones tended to report broader high-risk ranges [14,24,25]. Other patterns identified by non-linear modeling approaches (increasing and decreasing trends) may reflect either substantial uncertainty at temperature extremes [18,31,33] or constraints arising from analyses limited to narrow ranges of temperature (often due to the use of annual metrics) [17,34].

### Humidity

A total of 11 studies investigated the relationship with humidity, all of which used relative humidity as the primary metric (**Table 2**). Most focused on national or subnational regions in China, with one analyzing nationwide data from South Korea [16]. Among seven studies positing a non-linear relationship, five identified increasing SFTS risk up to approximately 75% relative humidity. Of these, two reported a notable decline beyond this threshold, indicating a right-skewed reversed U-shaped pattern [24,27]. The remaining three reported a continued upward trend across the entire humidity range examined [30,31,33], although estimates at higher humidity levels were highly uncertain due to sparse data above 75%. Of four studies assuming a linear relationship, three found a positive association with SFTS incidence [15,20,22], while the study in South Korea reported a negative association (but not statistically significant at the 0.05 level) that was derived based on annual mean humidity data restricted to a narrow range of 66–69% [16].

### Precipitation

Fourteen studies explored the relationship with precipitation, with nine focusing on cumulative precipitation and five on mean values (**Table 3**). Studies applying non-linear models to cumulative precipitation often reported right-skewed reverse U-shaped associations, characterized by a sharp increase in risk beyond a threshold (inflection point) followed by a sustained plateau and gradual decline [24,36]. While the overall pattern was similar, inflection points varied across studies, alongside substantial heterogeneities in precipitation metrics, including differences in spatial resolution (national vs. subnational) and temporal scale (e.g., cumulative precipitation during the driest quarter [25]). Such inconsistencies complicate cross-study comparisons, particularly between subtropical and temperate zones, where precipitation regimes differ markedly [37]. Studies using mean precipitation measures lacked sufficient spatial and temporal granularity, limiting the ability to draw generalized conclusions [20,22,26,32,34].

### Elevation

Among nine studies examining the relationship with elevation (i.e., the height above mean sea level), all but one (which assumed a linear relationship [21]) reported a reverse U-shaped pattern (**Table S5**). Across these studies, low-to-moderate elevations were consistently associated with the highest SFTS risk, forming a right-skewed reverse U-shaped pattern, although the absolute magnitude of elevation varied across studies. Specifically, studies in China typically reported high-risk ranges between 100–400 meters, corresponding to hilly (<200 meters) and low-mountain (200–500 meters) zones based on their national topographic classification [38]. In contrast, a nationwide study from Japan identified peak SFTS hospitalization risk at much lower elevations (9.8–29.7 meters; classified as mid-elevation within that study), likely reflecting the overall lower elevation profile of the study regions, where the maximum elevation analyzed was 114 meters [18].

### Land cover

Twelve studies examined the relationship between land cover and SFTS occurrence (**Table S6**), with the majority (9 of 12) accounting for elevation as a potential confounder, given that higher elevations generally correspond to mountainous areas. Metrics for land cover varied across studies: three studies employed the normalized difference vegetation index (NDVI, ranging from -1 to 1), while the remaining nine used the percentage coverage of specific land types. Among the NDVI-based studies, two conducted in a temperate zone (Shandong) identified a reverse U-shaped pattern, with SFTS risk peaking in moderately vegetated areas (NDVI range of 0.3–0.75) and declining in more densely vegetated areas [34,36]. In contrast, a study from a subtropical zone (Zhejiang) reported a monotonically decreasing trend, with the high-risk range spanning a broad NDVI range (0–0.75) [17]. Among studies using specific land types, heterogeneity in the categories considered (e.g., forest, shrubland, grassland, cropland, farmland, water bodies) limited direct cross-study comparisons.

### Other environmental factors

The roles of sunshine duration, wind speed, and air pressure in SFTS outcomes have been explored in a limited number of studies, with no more than six examining each variable. Additionally, substantial heterogeneity in measurement approaches further complicates cross-study comparisons and synthesis (**S7–S9 Tables**). Given these, the current evidence remains insufficient to draw definitive conclusions regarding their influence on SFTS outcomes.

## Discussion

Environmental factors shape the transmission dynamics of vector-borne diseases, determining when and where transmission occurs. This role is particularly pronounced in the case of SFTS, where transmission to humans arises from a multi-host spillover system in which environmental conditions simultaneously affect tick dynamics, animal reservoir activity, and human exposure. To characterize these relationships, our systematic review of 2,910 studies synthesizes evidence across multiple geographic settings, measurement scales, and analytical frameworks. Despite considerable heterogeneity in study designs, environmental factors—including temperature, humidity, precipitation, elevation, and land cover—consistently exhibited non-linear associations with human SFTS occurrence, most commonly taking the form of reverse U-shaped patterns. These findings reinforce the necessity of analytic frameworks that can accommodate non-linear dynamics when inferring environmental effects that shape SFTS transmission risk. However, we found no quantitative assessment of how environmental factors influence SFTS occurrences in the dominant tick vector (*H. longicornis*) or animal reservoirs, despite such studies falling within our search strategy and thus expected to be captured. This absence reveals a critical blind spot, emphasizing the need for future research that integrates multi-host surveillance beyond human outcomes to better elucidate how environmental drivers shape SFTSV ecology at the tick–animal–human interface.

These reverse U-shaped relationships are best understood within a broader One Health perspective, where SFTS occurrence in humans serves as a downstream reflection of multi-layered environmental effects cascading through transboundary interactions among ticks, animal reservoirs, and human populations [39]. Such non-linear patterns likely arise through at least three major pathways: (i) regulation of tick population dynamics, (ii) modulation of viral circulation within vector populations, and (iii) shift in human and animal reservoir behaviors that affect spillover risk. First, environmental conditions—particularly temperature and humidity—play a central role in shaping the population dynamics and behavior of *H. longicornis*. While this species tolerates a broad thermal gradient [40], environmental extremes at either end disrupt its development, questing activity, and survival. High temperatures delay molting and suppress questing activity, with sharp increases in mortality above 40°C [40]. Conversely, extreme cold elevates overwintering mortality and reduces egg hatchability, limiting population growth in the following season [41]. Similarly, low humidity increases larval desiccation, whereas excessive humidity (>85%) impairs questing activity and heightens vulnerability to microbial infections [33].

Second, environmental conditions also critically shape the mechanisms that sustain SFTSV circulation within tick populations. Accumulating evidence suggests that both horizontal (via co-feeding) and vertical (via transovarial pathways) transmission pathways play essential roles in maintaining viral persistence in tick populations [42], yet these mechanisms are highly sensitive to environmental stressors. Extreme temperature and humidity can impair tick survival, reproduction, and life cycle progression, thereby constraining vertical transmission through reduced oviposition or larval viability and limiting horizontal transmission by suppressing questing activity or increasing mortality. Additionally, extreme precipitation events (e.g., heavy rainfall or flooding) may physically displace ticks from their habitats or host [43], further destabilizing viral circulation within tick populations.

Lastly, environmental factors also influence human behavior and wildlife movements, changing the likelihood of tick exposure. Outdoor activities associated with higher tick exposure risks—such as farming, forestry, and recreation (e.g., hiking or camping) [44]—are more frequent under moderate weather conditions and decline during climatic extremes. The observed reverse U-shaped relationship with land cover (i.e., coverage of grassland) may similarly reflect such behavioral dynamics; although ticks often flourish in densely vegetated environments, limited human access in some of these may reduce spillover opportunities to humans.

However, translating these environmental associations into actionable control strategies requires an integrated perspective, as SFTSV transmission reflects the interplay of multiple environmental conditions rather than any single factor acting in isolation. Endemic hotspots of SFTS often emerge where favorable ecological conditions converge, offsetting constraints imposed by individual drivers. For instance, Gangwon province in South Korea, despite experiencing the nation’s coldest winter temperatures (–4–0°C on average [45]), consistently reports the highest human incidence of SFTS [46]. This paradoxical pattern likely stems from the region’s extensive mountainous terrain, which provides suitable habitats for ticks and wildlife reservoirs, enabling sustained spillover to humans even under otherwise limiting climatic conditions.

Beyond environmental drivers, human sociocultural practices also introduce an additional layer of complexity to SFTS transmission, leading to distinct epidemiological patterns even under comparable environmental conditions. While China and Japan exhibit similar seasonal trends in human SFTS incidence—peaking in early summer (May–June) before gradually declining into autumn—South Korea shows a distinct pattern, with cases rising in early summer but peaking sharply in October before dropping to near zero by November (**Figure S1**). Several factors may contribute to this divergence (e.g., differences in agricultural practices such as dual harvesting in some regions of China); however, the generally comparable environmental conditions across the three countries [47], which likely support similar tick ecology, suggest that sociocultural behaviors in humans may play a key role. In South Korea, outdoor activities surge during Korean Thanksgiving (Chuseok) in late September and early October, when many people visit ancestral graves located in vegetation-rich areas. This seasonal increase in outdoor exposure [48], coupled with heightened agricultural activity (e.g., wild food foraging [44]), likely elevates the risk of tick encounters, contributing to the delayed seasonal peak in case counts. Furthermore, sporadic human SFTSV infections have been linked to direct contact with infected livestock or companion animals, particularly through exposure to blood or body fluids, placing veterinarians and animal handlers at heightened risk [49].

While our systematic review shows that reverse U-shaped relationships are consistently reported across environmental factors, generalizing key features (such as high-risk ranges) remains challenging due to methodological heterogeneity. Much of this variation is driven by differences in spatiotemporal resolution in both environmental exposures and SFTS outcomes.

To compensate for low SFTS incidence (typically <10 cases per month at the regional level in South Korea and Japan [50,51]), studies often rely on coarser resolution to maintain statistical power. However, this introduces a critical trade-off, as such aggregations may obscure underlying environmental effects by artificially shifting or broadening estimated high-risk ranges. Indeed, studies employing coarse spatiotemporal resolutions tend to report lower and more dispersed high-risk temperature ranges compared to more granular analyses. This likely reflects the smoothing of fluctuations under coarse aggregation, which can distort the apparent shape of environmental effects.

Methodological heterogeneity is further compounded by differing modeling frameworks, particularly regarding how lagged effects are handled. Given the likely time delay between environmental exposure and human SFTS occurrence, incorporating lag structures within the model frameworks can fundamentally reshape the estimated exposure-response relationship, underscoring the importance of applying approaches that explicitly incorporate such time lags (e.g., distributed lag models). However, although five studies employed models with lag structure [22,27,29,30,35], the impact of these methodological differences was not clearly evident in our review, likely reflecting the abovementioned limitation of coarse data resolution. When the aggregation interval exceeds the duration of underlying time lags, the resulting smoothing averages out time-dependent signals. This attenuation masks the comparative performance of different modeling approaches. Hence, robust inference would require high-resolution data alongside application of modeling frameworks capable of capturing these time-dependent dynamics.

Beyond improving data resolutions, future research should move toward refined analytical approaches capable of disentangling stage-specific environmental effects across the SFTS transmission cycle. Although prior studies have advanced understanding of key environmental correlates, most have relied on analytic frameworks that link environmental variables solely to human SFTS outcomes—one of endpoints of a transboundary transmission process. Such approaches collapse multifaceted environmental effects into overly simplified estimates, obscuring how specific environmental drivers operate at different stages of SFTS transmission, from shaping tick population dynamics and viral persistence within vectors to modulating transmission among animal reservoirs. This simplification also complicates the interpretation of estimated time lags between environmental exposure and SFTS outcomes, as lag structures are likely to be context-specific and stage-dependent.

In this context, mechanistic (mathematical) modeling grounded in the One Health framework [52] offers a promising way forward. By explicitly incorporating environmental influences into the ecological dynamics of vectors and animal reservoirs, as well as the stage-specific transmission dynamics of SFTSV, these models can elucidate how environmental factors act at each stage of the transmission cycle across the tick–animal–human interface—an insight fundamentally inaccessible to conventional approaches that rely solely on simplified associations with human case counts. Moreover, incorporating human mobility into these frameworks can strengthen inference by partially addressing spatial misclassification inherent in residence-based incidence data, accounting for infections acquired outside of the residence region but recorded after they return and seek care in their home region. However, mechanistic models typically require assumptions on the underlying transmission dynamics, including the functional form of environmental effects, whereas statistical modeling and machine-learning approaches infer those associations directly from previously observed data. To reconcile these perspectives, the findings from this review bridge this gap by providing an empirical basis for informing plausible parameterization in mechanistic models, particularly in light of the non-linear relationships identified across environmental factors.

In parallel, mechanistic modeling efforts must be supported by robust empirical data that capture the broader ecological context of SFTSV transmission. This requires surveillance systems that extend beyond human cases to systematically collect serological and genomic data across vectors, animal reservoirs, and human populations. Such multi-host surveillance is essential for clarifying underlying patterns of host exposure, viral evolution, and interspecies transmission—core components of a One Health understanding of SFTS transmission dynamics. Furthermore, as the virus continues expanding its geographic range—demonstrated by the ongoing northward shift of SFTS endemic zones in Japan [53]—and as climate change and migratory bird movements increase the likelihood of long-distance tick dispersal [54,55], surveillance must also extend to regions not previously considered at risk. Regional monitoring will be critical for detecting emerging hotspots, tracking shifts in transmission dynamics (including changes in tick species composition or dominance [56]), and enhancing our understanding of how environmental and anthropogenic factors are shaping the evolving landscape of SFTSV transmission across the tick–animal–human interface.

Several limitations must be noted. First, a quantitative synthesis (e.g., meta-analysis) of evidence on environmental effects on SFTS occurrence was not conducted in this review, as substantial heterogeneity in analytical frameworks across studies precluded pooling reported outcomes on a common quantitative scale (including exposure-response curves or regression coefficients). Second, for similar reasons, standardized schematic diagrams for individual environmental factors were not presented, as variation in exposure definitions and measurement scales across studies would imply cross-study comparability that is not methodologically warranted. Third, such heterogeneity across studies further limited detailed cross-study comparisons, including the identification of potential climate zone-specific patterns in environmental effects on SFTS occurrence. As additional studies with more comparable conditions become available, more robust quantitative syntheses and cross-study comparisons may be feasible in future work. Lastly, although analytical frameworks beyond linearity assumptions may influence reported environmental effects, these differences were not systematically evaluated, as this was beyond the primary scope of the review.

## Conclusions

Our systematic review suggests that environmental factors are associated with human SFTS occurrence, most often through non-linear patterns. Beyond synthesizing existing evidence, we found that current evidence remains heavily focused on human outcomes while largely overlooking the multi-layered environmental influences that shape SFTSV transmission across vectors and animal reservoirs. This imbalance represents a critical gap from a One Health perspective, obscuring the way in which environmental drivers shape transmission dynamics across the tick–animal–human interface and limiting our understanding of SFTSV circulation as an integrated ecological process. As SFTS continues to expand beyond historically endemic areas, future research must adopt a more integrative paradigm by applying mechanistic modeling approaches integrated within One Health surveillance to disentangle stage-specific environmental effects across the transmission cycle. In parallel, strengthened multi-host surveillance and sustained scientific collaboration across regions and sectors will also be essential for enhancing inference on environmental effects and for supporting evidence-based control strategies to mitigate the growing transboundary burden of SFTS.

## Financial Support

This research was supported by the Japan Agency for Medical Research and Development (JP223fa627004); Japan Society for the Promotion of Science (JP25K18372 to AH); the British Academy (ODA Challenge-Oriented Research Grant to AH); Centers for Disease Control and Prevention (Safety and Healthcare Epidemiology Prevention Research Development, 200-2016-91781 to S-mJ); Singapore Ministry of Health (PREPARE-S1-2022-02 to S-mJ). The funders had no role in study design, data collection and narrative synthesis, decision to publish, or preparation of the manuscript.

## Data availability

This review was based exclusively on published literature.

## Conflict of Interest

The authors have no conflicts of interest to declare.

## Notes

### Competing Interest Statement

The authors have declared no competing interest.

### Funding Statement

This research was supported by the Japan Agency for Medical Research and Development (JP223fa627004). AH is supported by Grants-in-Aid for Scientific Research from the Japan Society for the Promotion of Science (JP25K18372) and the British Academy ODA Challenge-Oriented Research Grant. S-mJ is funded by the Centers for Disease Control and Prevention (Safety and Healthcare Epidemiology Prevention Research Development; 200-2016-91781). The funders had no role in study design, data collection and analysis, decision to publish, or preparation of the manuscript.

### Summary of Updates

The manuscript has been revised, particularly in the Discussion section, where additional paragraphs have been added.

