## Supplementary Materials for "Systematic review of environmental factors associated with severe fever with thrombocytopenia syndrome occurrences"

### **Appendix Contents**

**Section 1. Search Strategy**

- **Table S1.** Search terms for the bibliographic database

**Section 2. Study Selection**

- **Table S2.** Eligibility criteria for study inclusion

**Section 3. Quality Assessment**

- **Table S3.** Risk of bias assessment for included studies

**Section 4. Overview of Included Study Characteristics**

- **Table S4.** Summary of characteristics of studies included in the review

**Section 5. Narrative Synthesis of Additional Environmental Factors**

- **Table S5.** Summary of included studies examining the association between elevation and SFTS occurrence
- **Table S6.** Summary of included studies examining the association between land cover and SFTS occurrence
- **Table S7.** Summary of included studies examining the association between sunshine duration and SFTS occurrence
- **Table S8.** Summary of included studies examining the associations between air pressure and SFTS occurrence
- **Table S9.** Summary of included studies examining the associations between wind speed and SFTS occurrence

**Section 6. Seasonality of human SFTS incidence**

- **Figure S1.** Seasonality of confirmed severe fever with thrombocytopenia syndrome cases in China, Japan, and South Korea, 2016–2019

**Section 7. References**

**Section 1. Search Strategy**

**Table S1. Search terms for the bibliographic database**

| **Database** | **Search Terms** |
| --- | --- |
| *Embase* | ('severe fever with thrombocytopenia syndrome'/exp OR 'severe fever with thrombocytopenia syndrome' OR sfts OR sftsv) AND ('environment'/exp OR 'ecology'/exp OR 'meteorology'/exp OR 'meteorological concepts'/exp OR 'climate'/exp OR 'weather'/exp OR 'temperature'/exp OR 'rain'/exp OR 'humidity'/exp OR 'wind'/exp OR 'air pressure'/exp OR 'topography'/exp OR 'altitude'/exp OR 'sunlight'/exp OR 'radiation'/exp OR 'urbanization'/exp OR 'forest'/exp OR 'conservation of natural resources'/exp OR 'seasons'/exp OR environment* OR ecolog* OR meteorolog* OR climat* OR weather OR temperature OR precipitation OR rain* OR humidity OR evaporation OR wind OR pressure OR topograph* OR terrain OR elevat* OR altitude OR sun* OR solar OR radiation OR day* OR night* OR urban* OR deforest* OR forest* OR land* OR vegetation* OR crop* OR season*) |
| *Global Health* | ("severe fever with thrombocytopenia syndrome" OR SFTS OR SFTSV) AND (Environment* OR Ecolog* OR Meteorolog* OR Climat* OR Weather OR Temperature OR Precipitation OR Rain* OR Humidity OR Evaporation OR Wind OR Pressure OR Topograph* OR Terrain OR Elevat* OR Altitude OR Sun* OR Solar OR Radiation OR Day* OR Night* OR Urban* OR Deforest* OR Forest* OR Land* OR Vegetation* OR Crop* OR Season*) |
| *PubMed* | ("Severe fever with thrombocytopenia syndrome"[MeSH Terms] OR ("Severe fever with thrombocytopenia syndrome"[Title/Abstract] OR "SFTS"[Title/Abstract] OR "SFTSV"[Title/Abstract])) AND ("Environment"[MeSH Terms] OR "Ecology"[MeSH Terms] OR "Meteorology"[MeSH Terms] OR "Meteorological Concepts"[MeSH Terms] OR "Climate"[MeSH Terms] OR "Weather"[MeSH Terms] OR "Temperature"[MeSH Terms] OR "Rain"[MeSH Terms] OR "Humidity"[MeSH Terms] OR "Wind"[MeSH Terms] OR "Air Pressure"[MeSH Terms] OR "topography, medical"[MeSH Terms] OR "Altitude"[MeSH Terms] OR "Sunlight"[MeSH Terms] OR "Electromagnetic Radiation"[MeSH Terms] OR "Urbanization"[MeSH Terms] OR "Forests"[MeSH Terms] OR "Conservation of Natural Resources"[MeSH Terms] OR "Seasons"[MeSH Terms] OR ("environment*"[Title/Abstract] OR "ecolog*"[Title/Abstract] OR "meteorolog*"[Title/Abstract] OR "climat*"[Title/Abstract] OR "Weather"[Title/Abstract] OR "Temperature"[Title/Abstract] OR "Precipitation"[Title/Abstract] OR "rain*"[Title/Abstract] OR "Humidity"[Title/Abstract] OR "Evaporation"[Title/Abstract] OR "Wind"[Title/Abstract] OR "Pressure"[Title/Abstract] OR "topograph*"[Title/Abstract] OR "Terrain"[Title/Abstract] OR "elevat*"[Title/Abstract] OR "Altitude"[Title/Abstract] OR "sun"[Title/Abstract] OR "Solar"[Title/Abstract] OR "Radiation"[Title/Abstract] OR "day"[Title/Abstract] OR "night*"[Title/Abstract] OR "urban*"[Title/Abstract] OR "deforest*"[Title/Abstract] OR "forest*"[Title/Abstract] OR "land*"[Title/Abstract] OR "vegetation*"[Title/Abstract] OR "crop*"[Title/Abstract] OR "season*"[Title/Abstract])) |
| *Scopus* | ( ( INDEXTERMS ( "Severe fever with thrombocytopenia syndrome" ) ) OR ( TITLE-ABS-KEY ( "Severe fever with thrombocytopenia syndrome" OR sfts OR sftsv ) ) AND ( ( INDEXTERMS ( Environment ) OR INDEXTERMS ( Ecology ) OR INDEXTERMS ( Meteorology ) OR INDEXTERMS ( "Meteorological Concepts" ) OR INDEXTERMS ( Climate ) OR INDEXTERMS ( Weather ) OR INDEXTERMS ( Temperature ) OR INDEXTERMS ( Rain ) OR INDEXTERMS ( Humidity ) OR INDEXTERMS ( Wind ) OR INDEXTERMS ( "Air Pressure" ) OR INDEXTERMS ( "Topography" ) OR INDEXTERMS ( Altitude ) OR INDEXTERMS ( Sunlight ) OR INDEXTERMS ( "Electromagnetic Radiation" ) OR INDEXTERMS ( Urbanization ) OR INDEXTERMS ( Forest ) OR INDEXTERMS ( "Conservation of Natural Resources" ) OR INDEXTERMS ( Seasons ) ) OR ( Environment* OR Ecolog* OR Meteorolog* OR Climat* OR Weather OR Temperature OR Precipitation OR Rain* OR Humidity OR Evaporation OR Wind OR Pressure OR Topograph* OR Terrain OR Elevat* OR Altitude OR Sun* OR Solar OR Radiation OR Day* OR Night* OR Urban* OR Deforest* OR Forest* OR Land* OR Vegetation* OR Crop* OR Season* ) )) |
| *Web of Science* | (ALL=("Severe fever with thrombocytopenia syndrome" OR SFTS OR SFTSV)) AND ALL=(Environment* OR Ecolog* OR Meteorolog* OR Climat* OR Weather OR Temperature OR Precipitation OR Rain* OR Humidity OR Evaporation OR Wind OR Pressure OR Topograph* OR Terrain OR Elevat* OR Altitude OR Sun* OR Solar OR Radiation OR Day* OR Night* OR Urban* OR Deforest* OR Forest* OR Land* OR Vegetation* OR Crop* OR Season*) |

**Section 2. Study Selection**

Study selection eligibility criteria were structured across six key domains: publications, outcome, exposure, study design, analytic approach, and study focus. Since this review aims to synthesize evidence on the shape of relationships between environmental factors and SFTS outcomes, studies that did not provide any interpretable functional form of these relationships—including those limited to reporting simple correlation coefficients—were excluded. Details of each domain are provided below.

**Table S2. Eligibility criteria for study inclusion**

| **Criteria** | **Inclusion criteria** | **Exclusion criteria** |
| --- | --- | --- |
| Publications | Peer-reviewed original research articles  Studies published in English | Studies not published in English |
| Outcome | Studies with empirical SFTS outcome data across any of tick, animal, and human populations | Studies only focused on distribution of populations (e.g., tick population) |
| Exposure | Studies assessing relationship between environmental determinants and SFTS outcomes | Studies without assessment of environmental factors |
| Study design | Original quantitative studies | Clinical case reports, review articles, editorials, commentaries, conference abstracts, and qualitative research |
| Analytic approach | Studies using analytic methods to quantify associations between environmental factors and SFTS outcomes | Studies using descriptive statistics only (including simple correlation analysis without further analysis) |
| Study focus | Studies examining environmental factors in relation to SFTS outcomes | Studies focused exclusively on clinical management, laboratory diagnostics, pathogen characterization, or molecular studies unrelated to environmental analysis |

**Section 3. Quality Assessment**

Quality assessment was conducted using a modified version of the *Joanna Briggs Institute* (JBI) critical appraisal checklist for analytical cross-sectional studies, adapted to meet the requirements of environmental epidemiological research. Building on the original eight items (referred to as **Original Items**), we introduced modifications (referred to as **Modified Items**) to better capture the risk of biases related to variable selection and analytic approaches. Specifically, **Original Item 4**, “*Were objective, standard criteria used for measurement of the condition?*”, was divided into two items, one assessing environmental drivers (**Modified Item 4**) and the other assessing SFTS-related outcome (**Modified Item 7**), to allow for a more comprehensive evaluation of the standard criteria used to measure these components. In addition, **Original Item 5**, “*Were confounding factors identified?*”, was separated into two parts: “*Were confounding factors identified?*” (**Modified Item 5**), to evaluate whether relevant confounding variables were considered, and “*Were strategies to deal with confounding factors stated?*” (**Modified Item 6**), to assess whether environmental factors included in the analysis (including confounding variables) were supported by literature or domain knowledge, and whether statistical methods (e.g., Variance Inflation Factor for multicollinearity) were applied to address potential biases. Lastly, two new items were introduced to evaluate model selection and validation (**Modified Items 9 & 10**). “*Were model selection processes clearly stated*?” (**Modified Item 9**) examines whether model selection (i.e., comparing alternative models with different variable sets) was conducted systematically using evaluation measurements (e.g., area under the curve; AUC), while “*Were model validation processes clearly stated?*” (**Modified Item 10**) evaluates whether internal or external validation of the selected model was performed (e.g., cross-validation or spatial comparisons with empirical data from other regions). All other assessment criteria followed the standard JBI guidelines. Pairs of reviewers independently rated each domain as: yes (1), no (0), or unclear (0.5).

**Table S3. Assessment of risk of bias based on an adaptation of JBI Critical Appraisal Tool**

| **Author**  **(Year)** | **1. Were the criteria for inclusion in the sample clearly defined?** | **2. Were the study subjects and the setting described in detail?** | **3. Was the exposure measured in a valid and reliable way?** | **4. Were objective, standard criteria used for measurement of exposures?** | **5. Were confounding or key factors included?** | **6. Were strategies to deal with confounding factors and multicollinearity clearly stated?** | **7. Were objective, standard criteria used for measurement of the outcome?** | **8.Was appropriate statistical analysis used?** | **9. Were model selection processes clearly stated?** | **10. Were model validation processes clearly stated?** | **Overall**  **Score** |
| --- | --- | --- | --- | --- | --- | --- | --- | --- | --- | --- | --- |
| Du (2014) | 0.5 | 1 | 1 | 1 | 1 | 0 | 1 | 1 | 1 | 1 | 8.5 |
| Liu (2014) | 1 | 1 | 1 | 1 | 1 | 1 | 1 | 1 | 1 | 0 | 9 |
| Liu (2015) | 1 | 1 | 1 | 1 | 1 | 1 | 1 | 1 | 1 | 1 | 10 |
| Wu (2016) | 0.5 | 1 | 1 | 1 | 1 | 1 | 1 | 1 | 1 | 0 | 8.5 |
| Wang (2017) | 1 | 1 | 1 | 1 | 1 | 0 | 1 | 1 | 0.5 | 0 | 7.5 |
| Sun (2018a) | 1 | 1 | 1 | 1 | 1 | 0 | 1 | 1 | 1 | 1 | 9 |
| Sun (2018b) | 1 | 1 | 1 | 1 | 1 | 0 | 1 | 1 | 0 | 0 | 7 |
| Yasuo (2019) | 1 | 1 | 1 | 1 | 1 | 1 | 1 | 1 | 1 | 0.5 | 9.5 |
| Zhang (2019) | 0.5 | 1 | 1 | 1 | 1 | 1 | 1 | 1 | 1 | 1 | 9.5 |
| Miao (2020) | 0.5 | 1 | 1 | 1 | 1 | 1 | 1 | 1 | 1 | 0.5 | 9 |
| Wu (2020) | 1 | 1 | 1 | 1 | 1 | 0 | 1 | 1 | 0.5 | 0 | 7.5 |
| Miao (2021) | 1 | 1 | 1 | 1 | 1 | 1 | 1 | 1 | 1 | 1 | 10 |
| Sun (2021) | 1 | 1 | 1 | 1 | 1 | 1 | 1 | 1 | 0.5 | 0 | 8.5 |
| Deng (2022) | 1 | 1 | 1 | 1 | 1 | 1 | 1 | 1 | 1 | 1 | 10 |
| Jiang (2022) | 1 | 1 | 1 | 1 | 1 | 1 | 1 | 1 | 0 | 0 | 8 |
| Wang (2022) | 1 | 1 | 1 | 1 | 1 | 1 | 1 | 1 | 1 | 1 | 10 |
| Duan (2023) | 0.5 | 1 | 1 | 1 | 1 | 0 | 1 | 1 | 0.5 | 0.5 | 7.5 |
| Liu (2023) | 1 | 1 | 1 | 1 | 1 | 1 | 1 | 1 | 1 | 0 | 9 |
| Nam (2023) | 1 | 1 | 1 | 1 | 1 | 1 | 1 | 1 | 0.5 | 0 | 8.5 |
| Ding (2023) | 1 | 1 | 1 | 1 | 1 | 1 | 1 | 1 | 1 | 0.5 | 9.5 |
| Ogawa (2024) | 1 | 1 | 1 | 1 | 1 | 0 | 1 | 1 | 0 | 0 | 7 |
| Tao (2024) | 0.5 | 1 | 1 | 1 | 1 | 1 | 1 | 1 | 1 | 1 | 9.5 |
| Wang (2024) | 1 | 1 | 1 | 1 | 1 | 1 | 1 | 1 | 1 | 0 | 9 |

**Section 4. Overview of Included Study Characteristics**

**Table S4. Summary of characteristics of studies included in the review**

| **Author**  **(Year)** | **Country** | **Region (Climate zone†)** | **Study period** | **Model** | **Lag specification** | **SFTS outcome** | **Observational variables** |
| --- | --- | --- | --- | --- | --- | --- | --- |
| **Du (2014) [1]** | China | Shandong (Temperate) | 2010–2013 | MaxEnt | Not stated | Laboratory confirmed cases (timescale not reported) | Annual mean temperature, July mean temperature, January mean temperature, Annual mean relative humidity, January mean relative humidity, July mean relative humidity, Annual mean precipitation, January mean precipitation, July mean precipitation, Annual mean solar radiation hour, Annual mean air pressure, Annual mean vapor pressure, Land cover type, NDVI, Elevation |
| **Liu (2014) [2]** | China | Xinyang (Subtropical) | 2011–2012 | Poisson regression model | Not stated | Annual laboratory confirmed cases | Land cover types (irrigated cropland, rainfed cropland, orchard, forest, shrub, built-up land and water body), NDVI, Elevation, population density |
| **Liu (2015) [3]** | China | Nationwide | 2010–2013 | BRT coupled with MaxEnt | Lag examined, but not explictly reported | Monthly laboratory confirmed cases | Monthly mean temperature, monthly mean relative humidity, monthly cumulative precipitation, monthly cumulative sunshine hours, land cover (forest, shrub, cropland), elevation, cattle density, goat density, population density, distribution of tick (*H. longicornis* and *R. microplus*) |
| **Wu (2016) [4]** | China | Nationwide | 2010–2012 | GWLR | Not stated | Binary detection status based on laboratory confirmed cases (timescale not reported) | Monthly mean temperature, monthly mean precipitation, monthly mean relative humidity, mean proportion of rural population, mean proportion of primary industries from April to July over three years |
| **Wang (2017) [5]** | China | Hubei (Subtropical) | 2011–2016 | Panel Poisson regression model | Not stated | Annual laboratory confirmed & probable cases | Monthly temperature*, monthly humidity*, monthly precipitation*, % coverage of forest, irrigated cropland, rainfed cropland, grassland and built-up land, density of cattle, goat and human |
| **Sun (2018a) [6]** | China | Henan (Temperate) Hubei, Anhui (Subtropical) | 2011–2016 | Quasi-Poisson GAM | Not stated | Monthly laboratory confirmed cases | Monthly mean temperature, monthly maximum temperature, monthly minimum temperature, month mean relative humidity, monthly minimum relative humidity, monthly mean precipitation |
| **Sun (2018b) [7]** | China | Henan (Temperate) Hubei, Anhui (Subtropical) | 2011–2015 | Distributed lag nonlinear model | Cumulative lag effect (0–27 weeks) examined | Weekly laboratory confirmed cases | Weekly mean temperature, weekly maximum temperature, weekly minimum temperature, weekly mean relative humidity, weekly mean precipitation |
| **Yasuo (2019) [8]** | Japan | Miyazaki (Subtropical) | 2013–2018 | GWLR | Not stated | Annual laboratory confirmed cases | Annual mean temperature, annual mean humidity, % coverage of farmland, abandoned farmland and forest, elevation |
| **Zhang (2019) [9]** | China | Jiangsu (Subtropical) | 2010–2016 | MaxEnt | Not stated | Laboratory confirmed cases (timescale not reported) | Annual mean temperature, monthly mean temperature, mean temperature of driest quarter, maximum temperature of warmest month, temperature seasonality, mean diurnal range, annual cumulative precipitation, monthly mean precipitation, precipitation of driest month, precipitation of warmest quarter, NDVI, elevation, slope, aspect, monthly mean wind speed, solar radiation |
| **Miao (2020) [10]** | China, South Korea, and Japan | Nationwide | 2010–2018 | BRT coupled with MaxEnt | Not stated | Laboratory confirmed cases (timescale not reported) | Annual mean temperature, mean diurnal range, precipitation of warmest quarter, precipitation of coldest quarter, precipitation of driest month, % coverage of herbaceous vegetation of closed to open type, rainfed croplands, irrigated croplands, build-up land, water bodies, needle leaved forests, broadleaved forests, grasslands, other forests, and broadleaved deciduous forest of closed type, elevation, mammalian richness, livestock density, population density, predicted probability of H. longicornis presence |
| **Wu (2020) [11]** | China | Zhejiang (Subtropical) | 2011–2018 | Random forest model | Not stated | Monthly laboratory confirmed cases | Monthly mean temperature, monthly cumulative precipitation, monthly mean pressure, monthly mean relative humidity, monthly mean two-minute wind speed and monthly duration of sunshine |
| **Miao (2021) [12]** | China | Nationwide | 2010–2018 | Two–stage generalized BRT | Not stated | Annual laboratory confirmed cases | Annual mean temperature, mean diurnal range, temperature seasonality, maximum temperature of warmest month, minimum temperature of coldest month, annual range of temperature, mean temperature of wettest quarter, mean temperature of driest quarter, mean temperature of warmest quarter, mean temperature of coldest quarter, annual cumulative precipitation, precipitation of wettest month, precipitation of driest month, precipitation seasonality, precipitation of wettest quarter, precipitation of driest quarter, precipitation of warmest quarter, precipitation of coldest quarter, % coverage of cropland/shrub/woodland/etc, presence of tick species (e.g., H. longicornis), elevation, township-level and county-level socioeconomic data including GDP |
| **Sun (2021) [13]** | China | Nationwide | 2011–2018 | ENM with MaxEnt | Not stated | Annual laboratory confirmed cases | Annual mean temperature, annual mean relative humidity, annual cumulative precipitation, elevation, NDVI, land cover types, density of sheep, goat, cattle, poultry, and swine |
| **Deng (2022) [14]** | China | Jiangsu (Subtropical) | 2017–2020 | GAM | Lagged (30 days) and non-lagged effects examined | Daily laboratory confirmed cases, estimated tick–to–human transmissibility | Daily mean temperature, daily mean relative humidity, daily cumulative precipitation, daily mean wind speed, daily mean air pressure, daily mean sunshine duration, tick density |
| **Jiang (2022) [15]** | China | Shandong (Temperate) | 2010–2020 | GAM | Not stated | Total number of laboratory confirmed cases across the study period | Annual mean temperature, annual mean precipitation, % coverage of forest and grassland, NDVI, elevation, population density, meat production, milk production, GDP |
| **Wang (2022) [16]** | China | Liaoning (Temperate) | 2011–2019 | GLM | Lagged (1–5 weeks) and non-lagged effects examined | Monthly laboratory confirmed cases | Monthly mean temperature, monthly mean ground temperature, monthly mean precipitation, monthly mean relative humidity, monthly mean air pressure, monthly mean sunshine duration, monthly mean wind speed |
| **Duan (2023) [17]** | China | Shandong (Temperate) | 2010–2021 | MaxEnt | Not stated | Laboratory confirmed cases (timescale not reported) | Annual mean air temperature, annual mean relative humidity, annual mean wind speed, annual cumulative precipitation, NDVI, elevation, GDP, Nighttime light index, Density of chicken, cattle, goat and sheep |
| **Liu (2023) [18]** | China | Shangcheng (Subtropical) | 2011–2020 | NBR | Not stated | Annual laboratory confirmed cases | % coverages of cultivated land, forest, grassland, tea plantation, build-up land, bare land and water body, NDVI, elevation, density of human, swine, goat, poultry, distance to nearest large body of water, ratio (cultivated land area/forest land area), total incomes per person for each village |
| **Nam (2023) [19]** | South Korea | Nationwide | 2013–2018 | Bayesian regression model | Not stated | Annual laboratory confirmed & probable cases | Annual mean maximum temperature, annual relative humidity, annual cumulative precipitation, deforestation, urban area, population density, number of farmers, forest |
| **Ding (2023) [20]** | China | Nationwide | 2010-2019 | GAM | Not stated | Monthly laboratory confirmed cases | Monthly mean temperature, relative humidity, precipitation, % coverage of cropland, shrubland, grassland, forest |
| **Ogawa (2024) [21]** | Japan | Nationwide | 2010–2021 | Mixed–effects modified Poisson regression model | Not stated | Monthly hospitalizations | Monthly mean temperature, monthly cumulative precipitation, monthly mean solar radiation, elevation, type of vegetation (forest, farm or others) |
| **Tao (2024) [22]** | China | Zhejiang (Subtropical) | 2011–2019 | MaxEnt | Not stated | Laboratory confirmed & probable cases (timescale not reported) | Temperature seasonality, precipitation of wettest month, precipitation of wettest quarter, tick density, sunshine hours, mean relative humidity, mean land surface temperature, 20-8 precipitation, 8-20 precipitation, all-day precipitation, mean pressure, mean temperature, daily maximum temperature, mean wind speed, maximum wind speed. GDP, Digital elevation model, NDVI, density of cattle and sheep, EVI |
| **Wang (2024) [23]** | China | Jiaodong (Subtropical) | 2014–2020 | Quasi-Poisson GAM | Cumulative lag effect (0–27 weeks) examined | Weekly laboratory confirmed cases | Weekly lowest temperature, weekly mean temperature, weekly highest temperature, weekly mean air pressure, weekly mean sunshine duration, weekly mean wind speed, weekly mean speed of gustiness, maximum speed of gustiness, weekly mean relative humidity, weekly mean precipitation |

Abbreviations: MaxEnt, maximum entropy; BRT, boosted regression tree; GWLR, geographically weighted logistic regression; GAM, generalized additive model; ENM, ecological niche model; GLM, generalized linear model; NBR, negative binomial regression model.

† Climate zone was classified following the Trewartha climate classification.

* Monthly mean for the 4 months of the year when SFTS incidence rates are at their highest.

**Section 5. Narrative Synthesis of Additional Environmental Factors**

**Table S5. Summary of included studies examining the association between elevation and SFTS occurrence.**

| **Author**  **(Year)** | **Country** | **Region (Climate zone†)** | **Study period** | **Model** | **SFTS outcome** | **Observational variables** | **Range of the main variable (min–max)** | **Relationship with the main variable‡** | **Lag specification** |
| --- | --- | --- | --- | --- | --- | --- | --- | --- | --- |
| **Du (2014) [1]** | China | Shandong (Temperate) | 2010–2013 | MaxEnt | Laboratory confirmed cases (timescale not reported) | Elevation | -100–900m¶ | Reversed U–shape (high-risk range: 80–420 m) | Not stated |
| **Liu (2015) [3]** | China | Nationwide | 2010–2013 | BRT coupled with MaxEnt | Monthly laboratory confirmed cases | Elevation | 0–5000m¶ | Reversed U–shape (high-risk range: 100–400 m)* | Lag examined, but not explictly reported |
| **Yasuo (2019) [8]** | Japan | Miyazaki (Subtropical) | 2013–2018 | GWLR | Annual confirmed cases | Elevation | 3–1288m | Negative (not significant)§ | Not stated |
| **Miao (2020) [10]** | China, South Korea, and Japan | Nationwide | 2010–2018 | BRT coupled with MaxEnt | Laboratory confirmed cases (timescale not reported) | Elevation | 0–2500m¶ | Reversed U–shape (high-risk range: 0–1000 m) | Not stated |
| **Sun (2021) [13]** | China | Nationwide | 2011–2018 | ENM with MaxEnt | Annual laboratory confirmed cases | Elevation | -154–7518m | Reversed U–shape (high-risk point: 100m) | Not stated |
| **Miao (2021) [12]** | China | Nationwide | 2010–2018 | Two–stage generalized BRT | Annual laboratory confirmed cases | Elevation | 0–5000 m¶ | Reversed U-shape (high-risk point: 300m)* | Not stated |
| **Jiang (2022) [15]** | China | Shandong (Temperate) | 2010–2020 | GAM | Total number of laboratory confirmed cases across the study period | Elevation | 0–450 m¶ | Reversed U–shape (high-risk point: 300m) | Not stated |
| **Ogawa (2024) [21]** | Japan | Nationwide | 2010–2021 | Mixed–effects modified Poisson regression model | Monthly hospitalizations | Elevation | * Low: 1.4– 9.7m (mean: 4.7m)  * Middle: 9.8–29.7m (mean: 18.8m)  * High: 29.8–113.9m (mean: 56.1m) | Reversed U–shape** | Not stated |
| **Tao (2024) [22]** | China | Zhejiang (Subtropical) | 2011–2019 | MaxEnt | Laboratory confirmed & probable cases (timescale not reported) | Digital elevation model | 0–1918m | Reversed U–shape (high-risk range: 112–408 m) | Not stated |
| Abbreviations: MaxEnt, maximum entropy; GAM, generalized additive model; BRT, boosted regression tree; GWLR, geographically weighted logistic regression; NBR, negative binomial regression model.  † Climate zone was classified following the Trewartha climate classification. ‡ High-risk range is defined as either (1) conditions under which the maximum entropy model estimated an SFTS occurrence probability >50% or (2) the peak range reported by other models. When a range was not reported, the peak value is marked as the high-risk point. § A linear relationship between SFTS outcome and environmental variables was assumed. ¶ Range of the main variable was defined by the authors using the graph’s X-axis. * Range was defined by the reviewers through visual inspection of the original figures. ** Shape defined by the relationship of risk measures between categories of outcome variables, rather than in an exposure-response curve. | | | | | | | | | |

**Table S6. Summary of included studies examining the association between land cover and SFTS occurrence.**

| **Author**  **(Year)** | **Country** | **Region (Climate zone†)** | **Study period** | **Model** | **SFTS outcome** | **Observational variables** | **Range of the main variable (min–max)** | **Relationship with the main variable‡** | **Lag specification** |
| --- | --- | --- | --- | --- | --- | --- | --- | --- | --- |
| **Du (2014) [1]** | China | Shandong (Temperate) | 2010–2013 | MaxEnt | Laboratory confirmed cases (timescale not reported) | Monthly maximum NDVI | -0.1–1¶ | Reversed U–shape (high-risk range: 0.32–0.75) | Not stated |
| **Duan (2023) [17]** | China | Shandong (Temperate) | 2010–2021 | MaxEnt | Laboratory confirmed cases (timescale not reported) | NDVI | -0.1–1¶ | Reversed U–shape (high-risk range: 0.18–0.75) | Not stated |
| **Tao (2024) [22]** | China | Zhejiang (Subtropical) | 2011–2019 | MaxEnt | Laboratory confirmed & probable cases (timescale not reported) | NDVI | 0–1 | Decreasing shape. (high-risk range: 0–0.75)* | Not stated |
| **Jiang (2022) [15]** | China | Shandong (Temperate) | 2010–2020 | GAM | Total number of laboratory confirmed cases across the study period | % coverage of forest and grassland | 0–70%¶ | Reversed U–shape (high-risk point: 30%) | Not stated |
| **Liu (2014) [2]** | China | Xinyang (Subtropical) | 2011–2012 | Poisson regression model | Annual laboratory confirmed cases | % coverage of shrub, forest, rainfed cropland, quadratic rainfed cropland | Not reported | Shrub, forest, rainfed cropland: Positive (significant)§  Quadratic rainfed cropland: Negative (significant)§ | Not stated |
| **Liu (2015) [3]** | China | Nationwide | 2010–2013 | BRT coupled with MaxEnt | Monthly laboratory confirmed cases | % coverage of forest | 0–100%¶ | Constant shape with an initial increase at 10% (high-risk range: not defined) | Lag examined, but not explictly reported |
| **Wang (2017) [5]** | China | Hubei (Subtropical) | 2011–2016 | Panel Poisson regression model | Annual laboratory confirmed & probable cases | % coverage of built–up land, rainfed cropland | Not reported | Negative (significant)§ | Not stated |
| **Yasuo (2019) [8]** | Japan | Miyazaki (Subtropical) | 2013–2018 | GWLR | Laboratory confirmed cases | % coverage of farmland | 0–24.6% | Negative (not significant)§ | Not stated |
| **Miao (2020) [10]** | China, South Korea, Japan | Nationwide | 2010–2018 | BRT coupled with MaxEnt | Laboratory confirmed cases (timescale not reported) | % coverage of water bodies | 0–70% | Reversed U-shape (high-risk range: 0–25%) | Not stated |
| **Miao (2021) [12]** | China | Nationwide | 2010–2018 | Two–stage generalized BRT | Annual laboratory confirmed cases | % coverage of rainfed cropland, closed canopy woodland, shrubland, tea farm | Rainfed cropland: 0–95%* Closed-canopy woodland: 0–100%* Shrubland: 0–80%* Tea farm: 0–25%* | Rainfed cropland: reversed U- shape (high-risk point: 50%)* Closed canopy woodland and shrubland: increasing shape with a plateau at 5%* Tea farm: two-step increasing shape (high-risk point: 3%)* | Not stated |
| **Liu (2023) [18]** | China | Shangcheng (Subtropical) | 2011–2020 | NBR | Annual laboratory confirmed case | % coverage of forest land, tea plantation | Not reported | Positive (significant)§ | Not stated |
| **Ogawa (2024) [21]** | Japan | Nationwide | 2010–2021 | Mixed–effects modified Poisson regression model | Monthly hospitalizations | Type of vegetation (Forest, farm, others) | • High-level vegetation: forest • Mid-level vegetation: farm • Low-level vegetation: others | Increasing shape** | Not stated |
| Abbreviations: MaxEnt, maximum entropy; GAM, generalized additive model; BRT, boosted regression tree; GWLR, geographically weighted logistic regression; NBR, negative binomial regression model.  † Climate zone was classified following the Trewartha climate classification. ‡ High-risk range is defined as either (1) conditions under which the maximum entropy model estimated an SFTS occurrence probability >50% or (2) the peak range reported by other models. When a range was not reported, the peak value is marked as the high-risk point. § A linear relationship between SFTS outcome and environmental variables was assumed. ¶ Range of the main variable was defined by the authors using the graph’s X-axis. * Range was defined by the reviewers through visual inspection of the original figures. ** Shape defined by the relationship of risk measures between categories of outcome variables, rather than in an exposure-response curve. | | | | | | | | | |

**Table S7. Summary of included studies examining the association between sunshine duration and SFTS occurrence.**

| **Author**  **(Year)** | **Country** | **Region (Climate zone†)** | **Study period** | **Model** | **SFTS outcome** | **Observational variables** | **Range of the main variable (min–max)** | **Relationship with the main variable‡** | **Lag specification** |
| --- | --- | --- | --- | --- | --- | --- | --- | --- | --- |
| **Liu (2015) [3]** | China | Nationwide | 2010–2013 | BRT coupled with MaxEnt | Monthly laboratory confirmed cases | Monthly cumulative sunshine hours | 100–300h¶ | Reversed U–shape (high-risk range: 190–210h)* | Lag examined, but not explictly reported |
| **Wu (2020) [11]** | China | Zhejiang (Subtropical) | 2011–2018 | Random forest model | Monthly laboratory confirmed cases | Monthly cumulative sunshine hours | 41–275h (mean: 131.2h) | Increasing shape (high-risk range 209.7–275h) | Not stated |
| **Wang (2022) [16]** | China | Liaoning (Temperate) | 2011–2019 | GLM | Monthly laboratory confirmed cases | Monthly mean sunshine hours | Not reported (mean: 7.6h/day) | Direction varied across time lag (not significant)§ | Lagged (1–5 weeks) and non-lagged effects examined |
| **Wang (2024) [23]** | China | Jiaodong (Subtropical) | 2014–2020 | Quasi-Poisson GAM | Weekly laboratory confirmed cases | Weekly mean sunshine hours | 1.6–7.4h (mean: 4.4h) | Increasing shape (high-risk range: not defined) | Cumulative lag effect (0–27 weeks) examined |
| **Deng (2022) [14]** | China | Jiangsu (Subtropical) | 2017–2020 | GAM | Daily laboratory confirmed cases | Daily sunshine hours | 0–12h | Constant shape with an increase at 11h | Lagged (30 days) and non-lagged effects examined |
| **Ogawa (2024) [21]** | Japan | Nationwide | 2010–2021 | Mixed–effects modified Poisson regression model | Monthly hospitalizations | Monthly mean solar radiation | • Low: 11.0–12.8MJ/m^2^ (mean: 12.3MJ/m^2^) • Middle: 12.9–13.4MJ/m2 (mean: 13.2MJ/m^2^) • High: 13.5–15.9MJ/m^2^ (mean: 13.9MJ/m^2^) | Increasing shape** | Not stated |
| Abbreviations: BRT, boosted regression tree; MaxEnt, maximum entropy; GLM, generalized linear model; GAM, generalized additive model; h, hours.  † Climate zone was classified following the Trewartha climate classification. ‡ High-risk range is defined as either (1) conditions under which the maximum entropy model estimated an SFTS occurrence probability >50% or (2) the peak range reported by other models. When a range was not reported, the peak value is marked as the high-risk point. § A linear relationship between SFTS outcome and environmental variables was assumed. ¶ Range of the main variable was defined by the authors using the graph’s X-axis. * Range was defined by the reviewers through visual inspection of the original figures. ** Shape defined by the relationship of risk measures between categories of outcome variables, rather than in an exposure-response curve. *** All reported values were rounded to one decimal place. When results were reported as integers or presented graphically as ranges, values were shown as whole numbers. | | | | | | | | | |

**Table S8. Summary of included studies examining the associations between air pressure and SFTS occurrence.**

| **Author**  **(Year)** | **Country** | **Region (Climate zone†)** | **Study period** | **Model** | **SFTS outcome** | **Observational variables** | **Range of the main variable (min–max)** | **Relationship with the main variable‡** | **Lag specification** |
| --- | --- | --- | --- | --- | --- | --- | --- | --- | --- |
| **Wu (2020) [11]** | China | Zhejiang (Subtropical) | 2011–2018 | Random forest model | Monthly laboratory confirmed cases | Monthly mean air pressure | 999–1025hPa (mean: 1011.9hPa) | Declining shape (high-risk point: 1006.5hPa) | Not stated |
| **Wang (2022) [16]** | China | Liaoning (Temperate) | 2011–2019 | GLM | Monthly laboratory confirmed cases | Monthly mean air pressure | Not reported (mean: 992.2hPa) | Negative (significant) (0–5 week time lag)§ | Lagged (1–5 weeks) and non-lagged effects examined |
| Abbreviations: GLM, generalized linear model.  † Climate zone was classified following the Trewartha climate classification. ‡ High-risk range is defined as either (1) conditions under which the MaxEnt model estimated an SFTS occurrence probability >50% or (2) the peak range reported by other models. When a range was not reported, the peak value is marked as the high-risk point. § A linear relationship between SFTS outcome and environmental variables was assumed.  * All reported values were rounded to one decimal place. When results were reported as integers or presented graphically as ranges, values were shown as whole numbers. | | | | | | | | | |

**Table S9. Summary of included studies examining the associations between wind speed and SFTS occurrence.**

| **Author**  **(Year)** | **Country** | **Region (Climate zone†)** | **Study period** | **Model** | **SFTS outcome** | **Observational variables** | **Range of the main variable (min–max)** | **Relationship with the main variable‡** | **Lag specification** |
| --- | --- | --- | --- | --- | --- | --- | --- | --- | --- |
| **Duan (2023) [17]** | China | Shandong (Temperate) | 2010–2021 | MaxEnt | Laboratory confirmed cases  (timescale not reported) | Annual mean wind speed | 1.5–3.9m/s¶ | Reversed U–shape (high-risk range: 2.3–3.9 m/s) | Not stated |
| **Wu (2020) [11]** | China | Zhejiang (Subtropical) | 2011–2018 | Random forest model | Monthly laboratory confirmed cases | Monthly mean 2–minutes wind speed | 1–3m/s (mean: 1.8m/s) | Declining shape (high-risk range: 1.4–1.5m/s) | Not stated |
| **Wang (2022) [16]** | China | Liaoning (Temperate) | 2011–2019 | GLM | Monthly laboratory confirmed cases | Monthly mean wind speed | Not reported (mean: 2.4m/s) | Negative (significant) (0–5 week time lag)§ | Lagged (1–5 weeks) and non-lagged effects examined |
| **Wang (2024) [23]** | China | Jiaodong (Subtropical) | 2014–2020 | Quasi-Poisson GAM | Weekly laboratory confirmed cases | Weekly mean maximum speed of gustiness | 6–15.4m/s (mean:10.2m/s) | Constant shape (high-risk range: not defined) | Cumulative lag effect (0–27 weeks) examined |
| **Deng (2022) [14]** | China | Jiangsu (Subtropical) | 2017–2020 | GAM | Estimated tick–to–Human transmissibility | Daily mean wind speed | 0.7–6.3m/s | Constant shape (no time lag and 30-day time lag) | Lagged (30 days) and non-lagged effects examined |
| Abbreviations: MaxEnt, maximum entropy; GLM, generalized linear model; GAM, generalized additive model.  † Climate zone was classified following the Trewartha climate classification. ‡ High-risk range is defined as either (1) conditions under which the MaxEnt model estimated an SFTS occurrence probability >50% or (2) the peak range reported by other models. When a range was not reported, the peak value is marked as the high-risk point. § A linear relationship between SFTS outcome and environmental variables was assumed. ¶ Range of the main variable was defined by the authors using the graph’s X-axis.  * All reported values were rounded to one decimal place. When results were reported as integers or presented graphically as ranges, values were shown as whole numbers. | | | | | | | | | |

**Section 6. Seasonality of human SFTS incidence**

**Figure S1**. **Seasonality of confirmed severe fever with thrombocytopenia syndrome cases in China, Japan, and South Korea, 2016–2019**

Bars and numbers indicate monthly laboratory-confirmed human SFTS cases at the national level in (**A**) China, (**B**) Japan, and (**C**) South Korea from 2016–2019. Data were obtained from publicly available sources for China [24], Japan [25], and South Korea [26]. Dashed lines in each panel indicate the January 1st of each calendar year.

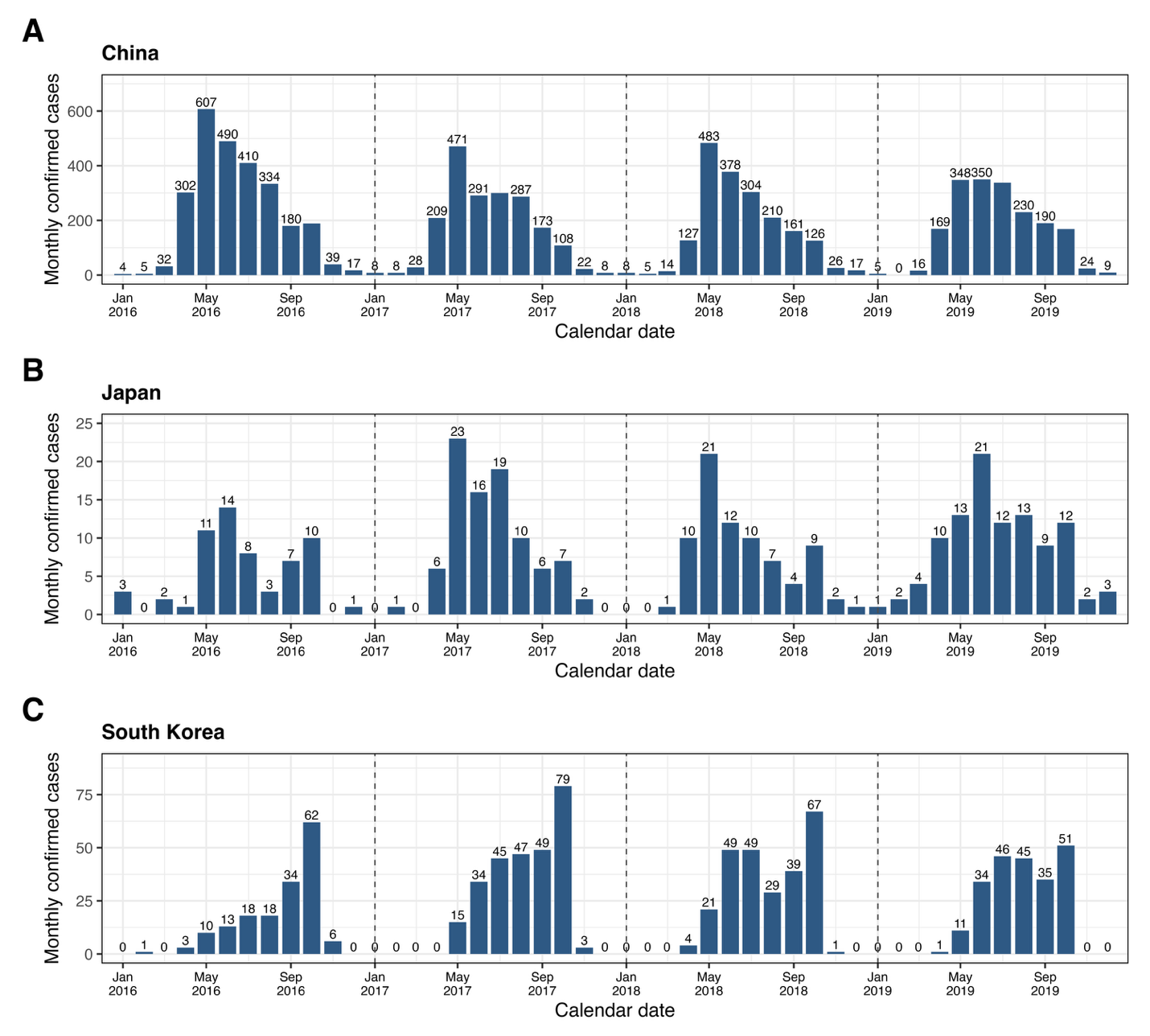
